# Repeated histological diagnoses and kidney graft failure: an observational cohort study

**DOI:** 10.64898/2026.02.17.26346474

**Authors:** Arthur Vranken, Maarten Coemans, Bertrand Chauveau, Tim Debyser, Priyanka Koshy, Dirk Kuypers, Christophe Masset, Angelica Pagliazzi, Thomas Vanhoutte, Karolien Wellekens, Thibaut Vaulet, Geert Verbeke, Maarten Naesens

**Author notes:** <u>Correspondence contact:</u> Name: Maarten Naesens, Address: Herestraat 49/7003, 3000 Leuven, Belgium.

## Abstract

**Background:** Banff histological diagnoses do not capture how a graft’s injury has evolved over time. Repeated injury is common, but the consequences of repeated histological injury are currently not known, especially when observed in the absence of clinical signs.

**Methods:** In a single-center observational cohort of 1818 kidney transplantations performed between 2004 and 2021, with 5218 post-transplant biopsies of which 82% were pre-planned protocol biopsies, transitions between six Banff histological diagnoses were described as recurrent events, with landmark Aalen-Johansen estimates and unadjusted Andersen-Gill rate ratios. A multistate model, with states for a first and a second diagnosis of rejection-related injury (RRI, comprising TCMR, AMR and MVI_DSA-/C4d-_), estimated cause-specific hazards of death-censored graft failure, in all biopsies and in protocol biopsies only. Follow-up ran from day 90 to five years after transplantation.

**Results:** Each diagnosis was more frequent after an earlier episode of the same diagnosis, and several diagnoses also made a different diagnosis more or less frequent: TCMR was more frequent after AMR, MVI_DSA-/C4d-_ and BKPyVAN, and AMR less frequent after MVI_DSA-/C4d-_. The hazard of graft failure was increased after a first RRI diagnosis (HR 2.18, 95% CI 1.35–3.51) and substantially further after a second (HR 9.31, 95% CI 5.84–14.85). The association persisted when both diagnoses were made on protocol biopsy, in the absence of clinical signs (HR 1.76, 95% CI 1.09–2.84 and HR 2.58, 95% CI 1.31–5.08), although the difference between a second and a first diagnosis was not statistically significant.

**Conclusions:** A prior Banff diagnosis strongly predicts that same diagnosis recurring, and in several cases also changes the expected incidence of a different diagnosis. A second diagnosis of rejection-related injury identifies a markedly higher-risk graft than a first, independently of baseline risk factors, and subclinical episodes detected on protocol biopsy also mark increased risk. This defines both an unmet therapeutic need and a high-risk population for trial enrichment.

**Key learning points:** *What was known:*

- A Banff diagnosis is rendered from a single biopsy, yet alloimmune injury often persists, recurs or changes over time.
- Repeated T-cell mediated rejection adds risk of graft failure, but evidence comes almost entirely from indication biopsies.
- How other diagnoses recur, and whether one predicts another, was largely unknown.

*This study adds:*

- In 1818 transplants with 5218 biopsies (82% protocol), a prior diagnosis strongly predicted its recurrence and altered the incidence of others.
- A second rejection-related episode carried a significantly higher hazard of death-censored graft failure than a first (9.31 versus 2.18).
- Subclinical protocol-biopsy episodes also carried risk (2.58 versus 1.76).

*Potential impact:*

- A first episode marks a graft at elevated risk, not a resolved problem. Biopsies are best read alongside the graft’s diagnostic history.
- Protocol biopsies detect prognostically relevant injury in apparently well recipients.
- Recipients with a second episode form a high-risk group suited to trials of more durable immune modulation.

## Introduction

Kidney transplantation is the preferred treatment for end-stage kidney disease, and immunosuppressive therapy has substantially reduced the incidence of early allograft rejection. Nonetheless, active and chronic injury remains prevalent and continues to drive premature return to dialysis.^1,2^ Such injury is most often detected histologically and categorized according to the Banff classification.^3^ A Banff diagnosis, however, is rendered from a single biopsy at a single moment. It is by construction mostly blind to the graft’s diagnostic history. Yet alloimmune injury is rarely a single event: it persists, is suppressed, recurs, and sometimes changes phenotype, and the same histological picture may mean something quite different in a graft seeing it for the first time than in one seeing it again.^4,5^

Evidence that this longitudinal context matters is accumulating but narrow. One study of repeated T-cell mediated rejection (TCMR) found that approximately half of patients with an initial TCMR experienced persistent or recurrent rejection, often within months, and that a second TCMR diagnosis carried a significant additional hazard of graft failure on top of the first.^6^ A recent multicenter study of early TCMR reported that, despite treatment, approximately one third of patients had rejection again on the first subsequent biopsy, and that these patients carried a substantially higher hazard of graft loss than those in whom rejection had resolved.^7^ Meanwhile the natural history of other phenotypes is contested: DSA-negative C4d-negative microvascular inflammation (MVI_DSA-/C4d-_) has been reported to be less often repeated,^8^ and BK polyomavirus nephropathy (BKPyVAN) is often regarded as manageable through reduction of immunosuppression without lasting consequence for the graft.^9,10^

Several questions therefore remain open. First, and most consequentially, the available evidence on repeated rejection rests on biopsies obtained because of graft dysfunction. Whether repeated injury detected *before* dysfunction, in a patient who appears entirely well, carries comparable prognostic weight has been examined only in small early cohorts,^11^ and has not been established in a contemporary adult population, because it requires a cohort with systematic sequential protocol biopsies. Second, existing work is largely confined to TCMR, leaving the recurrence behavior of antibody-mediated rejection (AMR), MVI_DSA-/C4d-_ and BKPyVAN, and the transitions between phenotypes, largely unexplored.

We therefore addressed two questions in an observational cohort with a systematic protocol biopsy program, in which the majority of biopsies were obtained irrespective of graft function. First, do prior Banff diagnoses predict which diagnosis follows, that is, do phenotypes persist or cluster, and at what rate? Second, does accumulating diagnoses of rejection-related injury carry risk beyond a first episode, and does that hold when the injury is detected on protocol biopsy in the absence of graft dysfunction?

## Methods

### Study population and clinical data

The cohort included all adult kidney transplant recipients transplanted at University Hospitals Leuven between 2004 and 2021 (clinicaltrials.gov ID NCT06505200). This is a single center operating a systematic protocol biopsy program, in which protocol biopsies are obtained at pre-specified time points in the absence of graft dysfunction and indication biopsies are obtained at times of graft dysfunction. Patients who underwent another solid organ transplantation before kidney transplantation, and patients with a combined transplantation, were not included.

Pre- and peri-operative clinical information is summarized in Table 1, and the complete adjustment set is given in the Supplementary Methods. Post-transplant clinical information consisted of anti-human leukocyte antigen donor-specific antibodies (HLA-DSA) and histological biopsy evaluation. Maintenance immunosuppression was mostly tacrolimus, mycophenolate and corticosteroids, with induction, usually basiliximab, reserved for recipients at increased immunological risk. From 2005, BK polyomavirus DNAemia led to mycophenolate reduction, and BKPyVAN to mycophenolate cessation, lower tacrolimus targets and cidofovir. Rejection treatment was at physician discretion, with steroid initiation recorded after each biopsy. Data collection and analysis were approved by the Ethics Committee of the University Hospitals Leuven (registration S64006). This paper adheres to the Declaration of Istanbul.

**Table 1.**
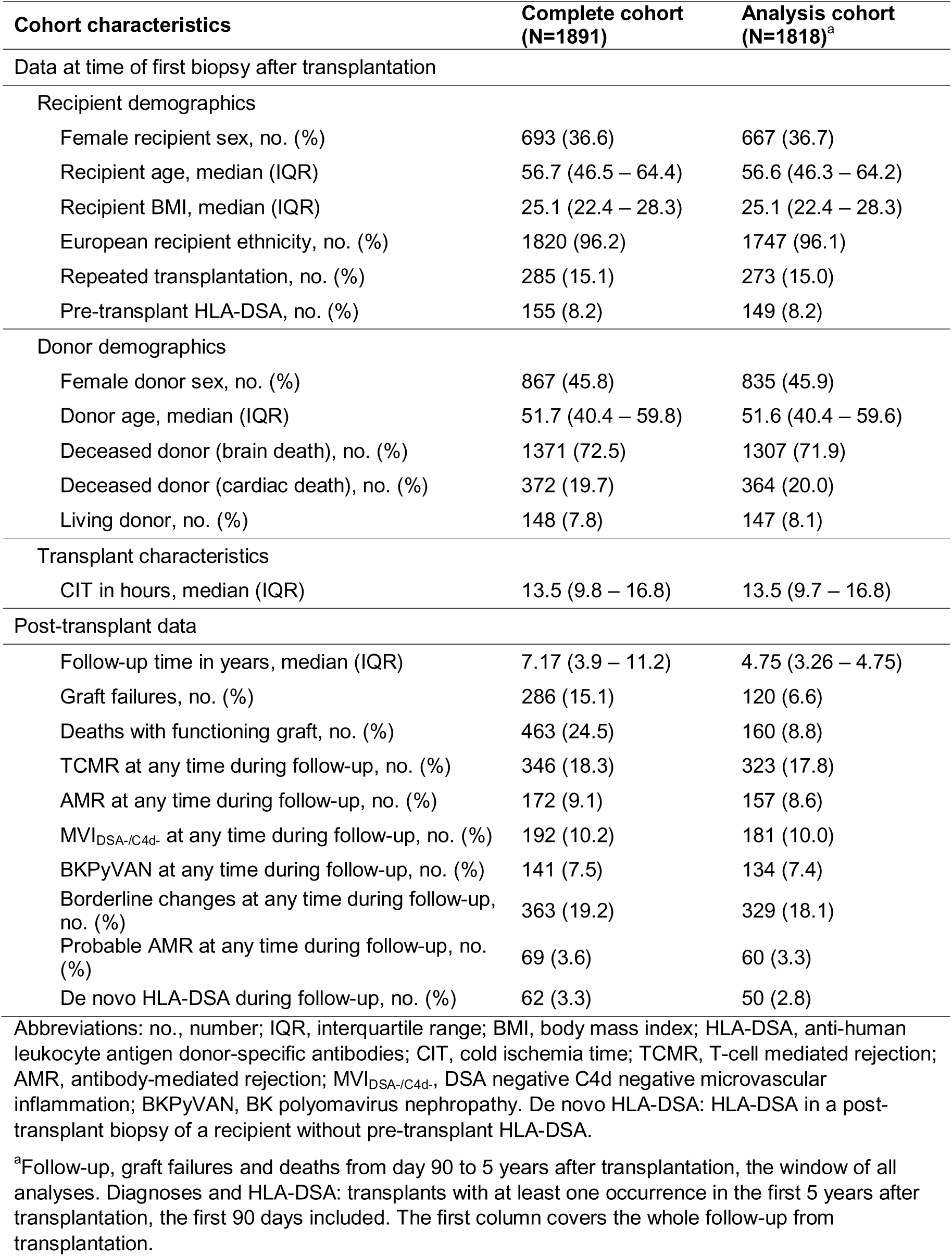
Kidney transplant characteristics.

### Kidney transplant biopsy evaluation

Post-transplantation indication biopsies were performed at the time of graft dysfunction, and pre-planned protocol biopsies were performed at 3, 12 and 24 months. The following, potentially co-existing, diagnoses were considered: TCMR, AMR, MVI_DSA-/C4d-_, BKPyVAN, Borderline changes (bTCMR) and Probable AMR (pAMR).^12^ Diagnoses were defined according to the Banff 2022 criteria,^3^ except for TCMR, from which isolated v cases were excluded.^13^

For the graft failure analyses, we defined a composite exposure, rejection-related injury (RRI), comprising TCMR, AMR and MVI_DSA-/C4d-_, phenotypes of established or suspected alloimmune origin that are each managed by escalating immunosuppression. bTCMR, pAMR and BKPyVAN were excluded from the composite and analyzed separately. The rationale for the composite boundaries is given in the Supplementary Methods. Whether the composite behaves consistently across its three components is examined directly (**Supplementary Figure S2**). "No Rejection/No BKPyVAN" refers to a biopsy in which none of the above diagnoses was present.

We used the term "repeated diagnosis" to refer to a repeated observation of the same diagnosis in two separate biopsies. We did not distinguish persistence from recurrence, since no systematic post-treatment biopsies were available. This distinction is likewise unresolved in the largest available cohort.^7^

### Statistics

Characteristics of transplants and follow-up intervals were summarized as medians and IQRs or counts and proportions. In all analyses, follow-up ran from day 90 to five years after transplantation, the period of biopsy collection, and the diagnostic history covered all biopsies of the first five years. Among recipients with multiple biopsies, all chronologically ordered within-patient biopsy pairs with the later biopsy after day 90 were classified by diagnostic sequence. Biopsies with concomitant diagnoses contributed to each applicable sequence: a pair of biopsies both showing concomitant TCMR and AMR (Mixed Rejection) contributed to the sequences TCMR-TCMR, TCMR-AMR, AMR-TCMR and AMR-AMR. These sequences were visualized in Sankey plots. This gives a general, time-agnostic description of transitions between diagnoses.

Transitions between diagnoses were further analyzed as recurrent events, since each diagnosis can be observed repeatedly and together with other diagnoses. Landmark Aalen-Johansen estimates gave the cumulative incidence of transitions into each later diagnosis from day 90, among transplants with a given diagnosis by day 90, with graft failure and death as competing events. These groups overlap and should not be interpreted as stratified cumulative incidences. Andersen-Gill^14^ models summarized each pair of earlier and later diagnosis in one number: the rate ratio of transitions into the later diagnosis, with the earlier diagnosis as the only time-dependent covariate. In a sensitivity analysis, steroid-treated and untreated earlier diagnoses were separated. All of these analyses are unadjusted for potential confounders and hence only describe the observed transitions.

A multistate model linked rejection-related injury to graft failure,^15^ with states 1) no RRI, 2) first RRI, 3) second RRI, 4) graft failure and 5) death with functioning graft (**Figure 1**). Landmark Aalen-Johansen estimates gave the cumulative incidence of graft failure from one year post-transplantation, by state at one year. A cause-specific Cox model for the transitions into graft failure, with the current RRI state as a time-dependent covariate, estimated the hazard ratios of a first and a second RRI. It was adjusted for baseline transplant characteristics and pre-transplant HLA-DSA, but not for potential mediators: time-dependent HLA-DSA (which also enters several diagnostic definitions), chronic lesion scores and the number of indication biopsies. Deviations from proportional hazards were assessed with the cox.zph function of the survival R package,^16^ and time-varying effects were modeled by an interaction with time since transplantation. A pre-specified secondary model counted only diagnoses made in protocol biopsies, thereby estimating the association in the absence of graft dysfunction and removing the dependence of exposure ascertainment on clinical presentation. In a sensitivity analysis, the exposure was restricted to steroid-treated episodes, with follow-up censored at the first RRI episode that was not steroid-treated.

**Figure 1.**
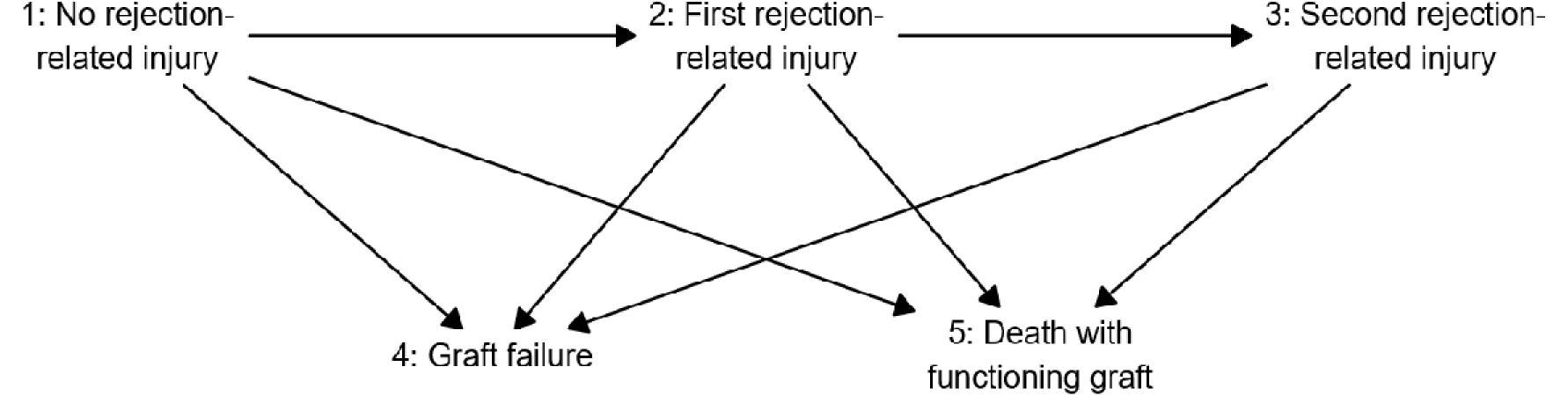
State space of the multistate model of rejection-related injury and graft failure. Diagram representing progression paths following kidney transplantation with regard to rejection-related injury, graft survival and patient survival.

Exploratory cause-specific Cox models, with the same adjustment set and follow-up, evaluated each individual diagnosis and each ordered pair of diagnoses as time-dependent covariates. All analyses were conducted in R version 4.4.1. Full model specification, the counting-process construction, censoring rules and the complete adjustment set are given in the Supplementary Methods.

## Results

### Study population and follow-up information

The complete cohort consisted of 1891 transplants. Characteristics are described in **Table 1**. Seventy-seven transplants had no biopsies, while the remaining 1814 transplants had 6272 biopsies in total, of which 4565 (73%) were protocol biopsies. After excluding cases with missing values or 90 days or less of follow-up (N = 73) from the complete cohort, 1818 transplants were included in all analyses. During follow-up (median 4.75 years, IQR 3.26–4.75), 120 grafts failed and 160 patients died with a functioning graft.

The analysis cohort contributed 5218 biopsies and 5900 time intervals: 682 (12%) intervals preceding the first post-transplantation biopsy, 4277 (72%) following a protocol biopsy and 941 (16%) following an indication biopsy (**Figure 2**). Their distribution over time, and of the time-dependent covariates within them, is summarized in **Supplementary Table S2**.

**Figure 2.**
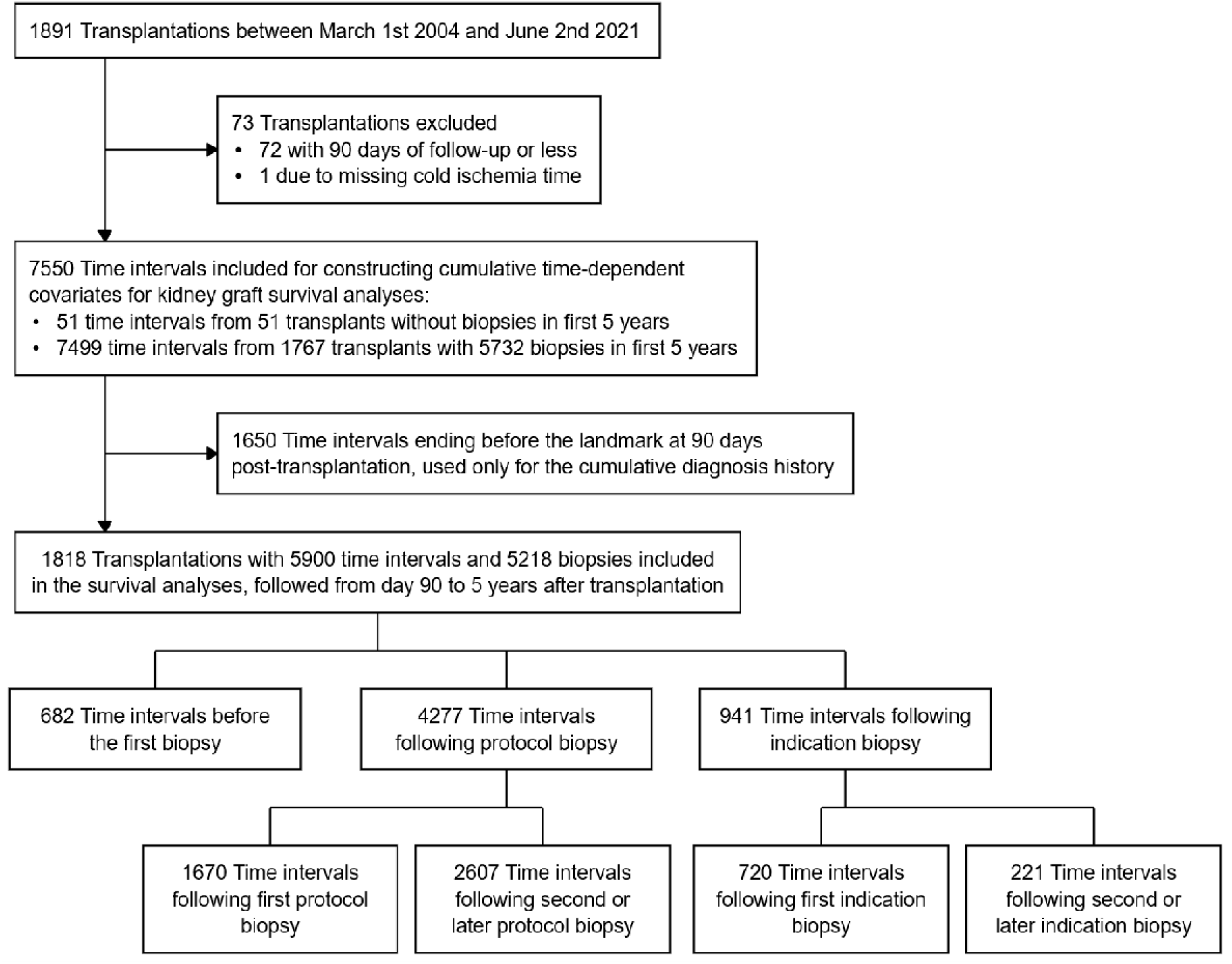
Flowchart for inclusion and exclusion of transplants and follow-up time intervals for survival analyses.

### Sequences of histological diagnoses

Of the 1818 analyzed transplants, 1562 (85.9%) had a biopsy after day 90 preceded by an earlier biopsy within five years, 309 (17.0%) had a diagnosis of rejection or of BKPyVAN in such a biopsy and in an earlier one, and 158 (8.7%) had such diagnoses in protocol biopsies specifically. Among these 1562 transplants, all sequences of diagnoses were counted and visualized (**Figure 3**, **Supplementary Table S3**). Among sequences between two diagnoses, repeated observation of the same diagnosis was more common than transition between different diagnoses. Heterogeneous transitions were still relatively common between TCMR and AMR and between TCMR and MVI_DSA-/C4d-_, while the sequence from MVI_DSA-/C4d-_ to AMR was very rare (**Figure 3**).

**Figure 3.**
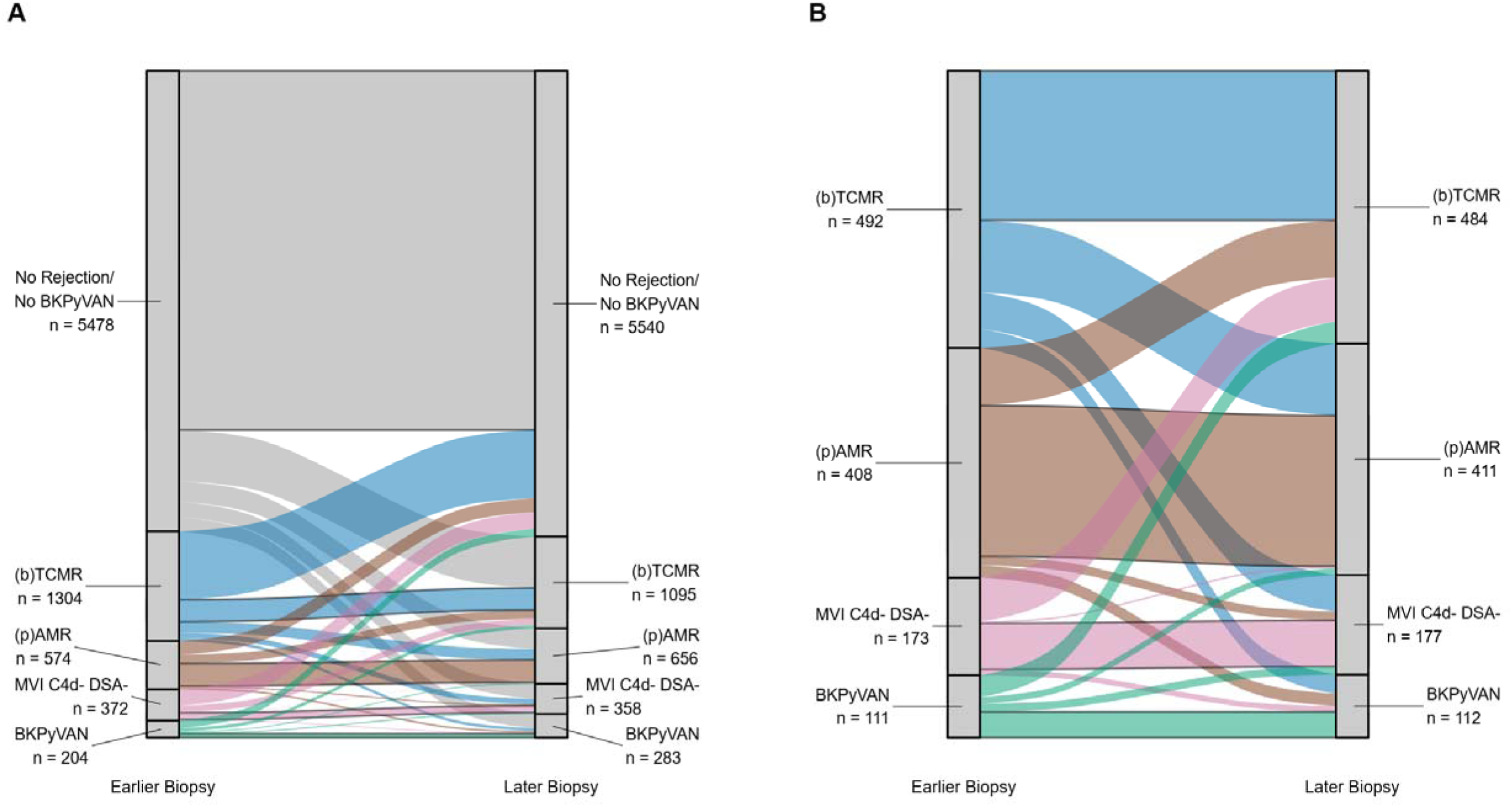
Sequences of diagnoses in pairs of biopsies within the first five years after transplantation, the later biopsy after day 90 (N=1562 transplants). All possible within-patient pairs of diagnoses were allocated to one of the Sankey flows, including pairs of diagnoses with other diagnoses in between. Biopsies with multiple simultaneous diagnoses (e.g. TCMR and AMR, also referred to as Mixed Rejection) contributed to the Sankey flows for each of those diagnoses. (A) Sequences including No Rejection/No BKPyVAN. (B) Sequences excluding No Rejection/No BKPyVAN. Abbreviations: (b)TCMR, T-cell mediated rejection or Borderline changes; (p)AMR, antibody-mediated rejection or probable antibody-mediated rejection; MVI_DSA-/C4d-_, DSA-negative C4d-negative microvascular inflammation; BKPyVAN, BK polyomavirus nephropathy.

Cumulative incidence curves from day 90 revealed heterogeneity in the timing of repeated diagnoses. Repeated BKPyVAN occurred mostly within weeks after day 90, whereas repeated AMR and MVI_DSA-/C4d-_ also accrued around the one-year protocol biopsy, after which the curves flattened (**Figure 4**).

**Figure 4.**
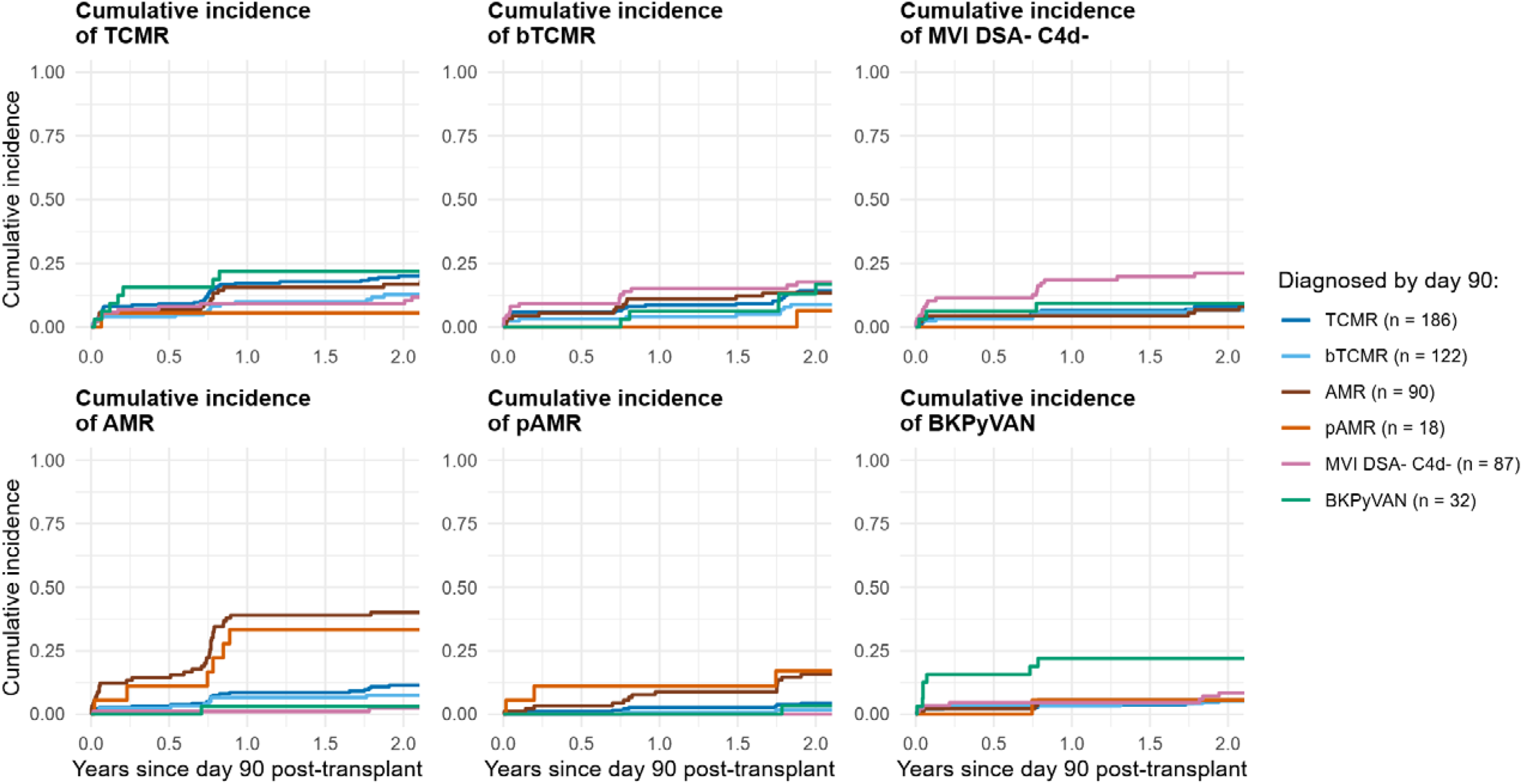
Cumulative incidence of transitions into each later diagnosis after day 90, by diagnosis made by day 90 (N=1818 transplants, with group sizes shown in the figure legend). Curves are landmark Aalen-Johansen estimates from day 90 after transplantation, with graft failure and death as competing events and follow-up censored at five years after transplantation. Groups overlap, since a transplant can have several diagnoses by day 90. Curves are shown up to two years after day 90 and stop once fewer than 10 transplants remain at risk. Abbreviations: TCMR, T-cell mediated rejection; bTCMR, Borderline changes; AMR, antibody-mediated rejection; pAMR, Probable AMR; MVI_DSA-/C4d-_, DSA-negative C4d-negative microvascular inflammation; BKPyVAN, BK polyomavirus nephropathy

Andersen-Gill rate ratios, which summarize each pair of diagnoses in one number, confirmed the visual impressions from the Sankey plots and the cumulative incidences. The rate of transitions into every diagnosis was significantly increased after an earlier diagnosis of the same type (RR 1.49 for bTCMR to 23.67 for AMR). For every diagnosis except bTCMR, this was the largest rate ratio of any earlier diagnosis. Several transitions between different diagnoses were also more frequent: TCMR after AMR, MVI_DSA-/C4d-_ and BKPyVAN, AMR after TCMR, bTCMR and pAMR, and bTCMR and pAMR after AMR. AMR was less frequent after MVI_DSA-/C4d-_ (RR 0.20) and BKPyVAN after bTCMR (RR 0.42) (**Figure 5**, **Supplementary Table S4**). In the sensitivity analysis of steroid-treated earlier diagnoses, TCMR, AMR, pAMR and MVI_DSA-/C4d-_still occurred significantly more often after a prior diagnosis of the same type, though not to the same degree as in the main analysis, especially for AMR, pAMR and MVI_DSA-/C4d-_(**Supplementary Figure S1**, **Supplementary Table S5**).

**Figure 5.**
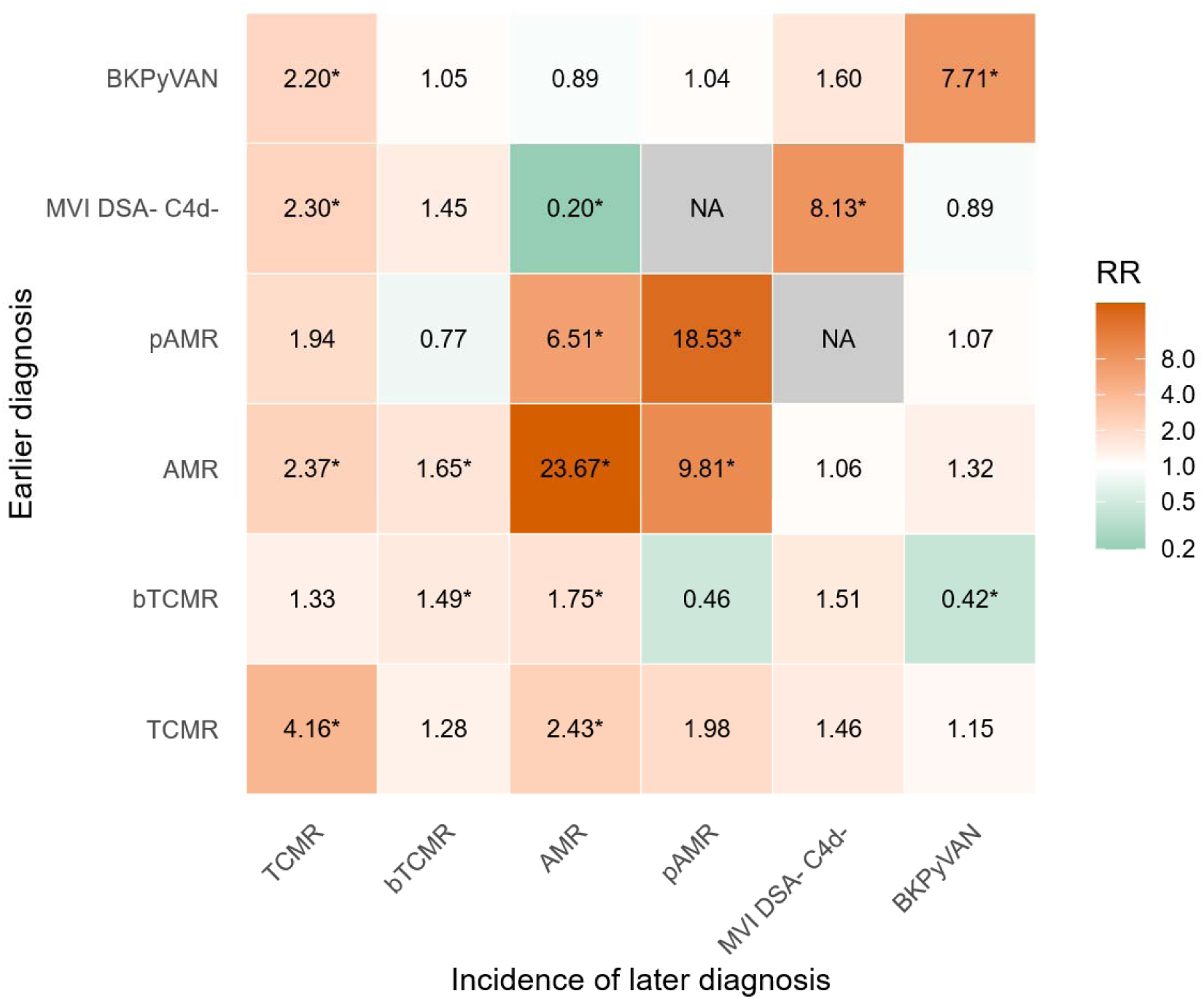
Rate ratios of transitions into later diagnoses after earlier diagnoses (N=1818 transplants). Each rate ratio is obtained from a separate unadjusted Andersen-Gill model for transitions into the later diagnosis, with the earlier diagnosis as its only covariate, with follow-up from day 90 and censored at five years after transplantation. Abbreviations: RR, rate ratio; NA, not applicable due to lack of diagnoses; TCMR, T-cell mediated rejection; bTCMR, Borderline changes; AMR, antibody-mediated rejection; pAMR, Probable AMR; MVI_DSA-/C4d-_, DSA-negative C4d-negative microvascular inflammation; BKPyVAN, BK polyomavirus nephropathy. *P < .05

### Associations with graft failure

#### Repeated rejection-related injury and graft failure

From the one-year landmark to five years after transplantation, the cumulative incidence of graft failure was 4.3% without RRI, 7.1% after a first RRI diagnosis and 32.4% after a second (**Figure 6**). The hazard of graft failure was significantly increased after a first RRI diagnosis (HR 2.18, 95% CI 1.35–3.51) and markedly further after a second (HR 9.31, 95% CI 5.84–14.85). The effect of the second was significantly larger than that of the first (**Figure 6**, **Table 2**, **Supplementary Tables S6 and S9**). The proportional hazards test was borderline non-significant (P = .061). In a model allowing time-varying effects, the hazard ratio of a first RRI declined from 3.35 (95% CI 1.83–6.11) at one year to 0.70 (95% CI 0.21–2.28) at five years, whereas that of a second remained high, from 9.65 (95% CI 4.66–19.95) to 7.94 (95% CI 3.55–17.78) (**Supplementary Figure S3**).

**Figure 6.**
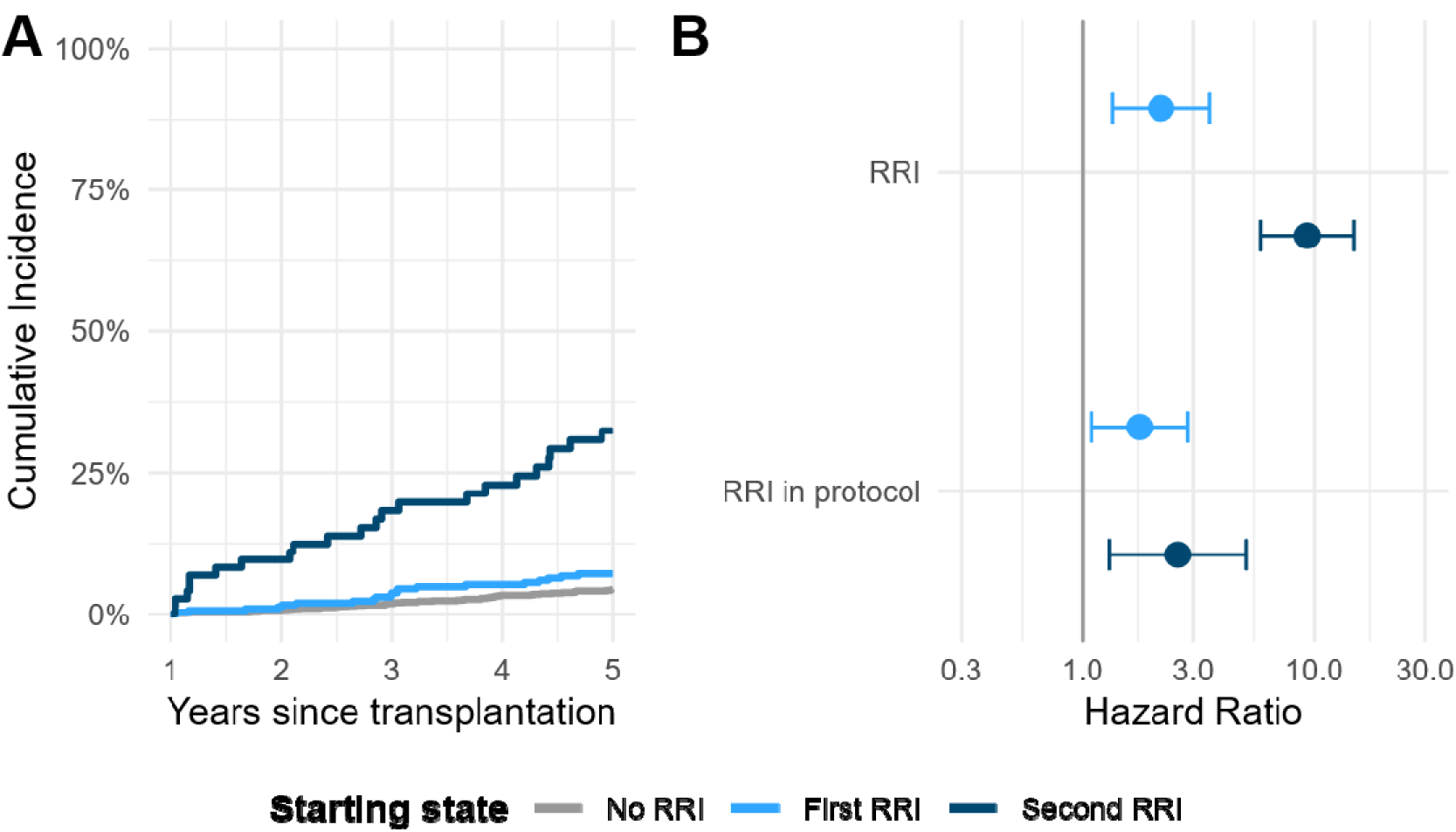
Cumulative incidences and cause-specific hazard ratios for kidney graft failure by history of rejection-related injury (RRI) (N=1818). (A) Cumulative incidences are landmark Aalen-Johansen estimates from one year after transplantation, in the transplants with no rejection-related injury (n=1383), one episode (n=309) or two or more episodes (n=73) at one year, with death with a functioning graft as a competing event. Follow-up was censored at five years after transplantation. (B) Each pair of cause-specific hazard ratios belongs to a separate model and is adjusted for clinical characteristics at time of transplantation and pre-transplant HLA-DSA status.

**Table 2.** Associations of a first and second diagnosis of rejection-related injury with the cause-specific hazard of graft failure.

| **Covariate** | **Multivariable HR\*** | **95% CI** | **P value** |
|---|---|---|---|
| In all biopsies |  |  |  |
| No rejection-related injury | *reference* | . | . |
| First rejection-related injury | 2.18 | 1.35 - 3.51 | 0.0014 |
| Second rejection-related injury | 9.31 | 5.84 - 14.85 | <.0001 |
| In protocol biopsies<sup>a</sup> |  |  |  |
| No rejection-related injury | *reference* | . | . |
| First rejection-related injury | 1.76 | 1.09 - 2.84 | 0.0202 |
| Second rejection-related injury | 2.58 | 1.31 - 5.08 | 0.0060 |

\*Models adjusted for repeat transplantation, donor status (living, brain death, cardiac death), male recipient sex, donor sex, recipient age, donor age, recipient ethnicity, recipient BMI, cold ischemia time, calendar year of transplantation and pre-transplant donor-specific anti-human leukocyte antigen antibodies (HLA-DSA). Follow-up was censored at five years after transplantation.
<sup>a</sup>Start-stop counting process format was reformulated on protocol biopsies only.
Note: complete model results are shown in Supplementary Table S6.
Abbreviations: HR, hazard ratio; CI, confidence interval.

#### Rejection-related injury detected on protocol biopsy

Restricting the exposure to diagnoses made in protocol biopsies, that is, to injury detected in the absence of graft dysfunction, the hazard of graft failure remained significantly increased after a first RRI diagnosis (HR 1.76, 95% CI 1.09–2.84) and after a second (HR 2.58, 95% CI 1.31–5.08), although the difference between first and second was not statistically significant (HR for second versus first 1.47, 95% CI 0.72–2.98) (**Figure 6**, **Table 2**, **Supplementary Tables S6 and S9**).

In the sensitivity analysis restricted to steroid-treated episodes, the conclusions were unchanged: both a first (HR 2.02, 95% CI 1.10–3.72) and a second RRI (HR 12.13, 95% CI 5.55–26.49) were associated with graft failure, the second with the larger hazard ratio (**Supplementary Table S7**).

#### Specific diagnoses

Examining diagnoses individually, the hazard of graft failure was significantly increased after each diagnosis except pAMR (HR 1.32, 95% CI 0.60–2.93). Each of the three components of the RRI composite was individually associated with an increased hazard of graft failure, indicating that the composite result is not attributable to a single constituent phenotype. The hazard of graft failure was also significantly increased by the majority of sequences of two diagnoses, and often more so than after a single diagnosis (**Supplementary Figure S2**, **Supplementary Table S8**).

## Discussion

In this cohort, a prior diagnosis was associated with a higher incidence of the same diagnosis later, and in some cases with a higher or lower incidence of a different phenotype, for instance more TCMR after AMR, MVI_DSA-/C4d-_ or BKPyVAN and less AMR after MVI_DSA-/C4d-_. The hazard of graft failure rose after a first episode of rejection-related injury and steeply after a second. Both episodes also carried risk when detected on protocol biopsy in patients without graft dysfunction, although the difference between them was not significant there. While the Banff classification mostly does not take previous biopsies into account, these findings indicate that the diagnostic history of a graft carries prognostic information that a single biopsy cannot convey.

Diagnoses that caused graft dysfunction are likely more severe, and so more consequential for graft survival, than those detected on protocol biopsy. This fits the attenuation of the hazard ratio of a second episode from 9.31 to 2.58 when only protocol biopsies were counted, and may explain part of the excess of a second over a first episode. Detection bias is the alternative: transplants that accumulate diagnoses are those biopsied most often, and indication biopsies are prompted by graft dysfunction. However, graft dysfunction can often be explained by histological injury diagnosed in the subsequent biopsy, so we believe detection bias to have limited impact. The protocol-biopsy model also addresses this concern by construction, since ascertainment then does not depend on clinical presentation. Both associations persisted there (HR 1.76 and 2.58), although the excess of a second over a first episode was no longer significant.

TCMR was often followed by a biopsy without rejection. Despite this, its association with graft failure was similar to that of AMR and MVI_DSA-/C4d-_. Established therapies for TCMR therefore appear effective at halting visible activity without fully curbing the effect on graft survival. This effect was especially pronounced when TCMR followed a prior diagnosis, confirming earlier findings.^6,7^

AMR recurred more strongly than any other diagnosis (RR 23.67) and was also frequent after pAMR (RR 6.51), plausibly reflecting persistent sensitization and the absence of adequate options to eliminate HLA-DSA.^17,18^ External evidence points the same way: after AMR treatment, persistence or progression of humoral lesions on a follow-up biopsy identified grafts at markedly higher risk of loss, whereas their disappearance was associated with more favorable long-term survival.^19^ A recent report likewise found an increased risk of AMR after pAMR, but also after MVI_DSA-/C4d-_, whereas in our study AMR was less frequent after MVI_DSA-/C4d-_ (RR 0.20).^20^ In our cohort we observed very little de novo occurrence of HLA-DSA after MVI_DSA-/C4d-_, which is the step that would be required for progression to AMR.

MVI_DSA-/C4d-_ has been described as more transient than AMR.^8^ Here it nevertheless recurred strongly (RR 8.13), and its association with graft failure was similar to that of AMR. This agrees with studies comparing the two in survival models using time of biopsy as time zero^20,21^ and in joint longitudinal-survival models.^22^ It differs from the weaker association reported by the same study that described the more transient course,^8^ which used a landmarked survival model and would therefore be expected to yield smaller effect sizes with reduced power.^23^ For MVI persisting across repeated biopsies, graft survival did not differ according to whether inflammation was still present on a third biopsy,^24^ although that design conditions on two prior episodes and so does not speak to the contrast between a first and a second diagnosis.

While BKPyVAN is held responsible for 5% to 10% of graft failures, previous studies have not found an effect on the hazard of graft failure,^25^ and BK polyomavirus can often be cleared by lowering immunosuppression.^10^ We nonetheless found that the incidence of BKPyVAN was increased after prior BKPyVAN, and that BKPyVAN was associated with an increased hazard of graft failure, especially after prior TCMR, pAMR or BKPyVAN. An increased incidence of TCMR after BK polyomavirus has also been reported in an independent cohort, plausibly reflecting the reduction of immunosuppression used to control viral replication.^26^ Repeated observation of BKPyVAN may mark more severe viral infection: earlier work associated repeated BKPyVAN with worsening eGFR trajectory^27^ and higher plasma viral load and histological pvl score with slower viral clearance.^25^

The principal strength of this study is the ability to analyze repeated diagnoses in protocol biopsies specifically, which allows the association to be assessed in the absence of a clinical indication. A second strength is the use of time-dependent covariates in recurrent-event and multistate models, which avoids immortal time bias arising from subgroups defined by time-dependent variables,^28^ handles correlated observations without discarding biopsies, and avoids the loss of power associated with landmarking.^23^ The Aalen-Johansen curves add the absolute-risk view relevant to stratification and trial enrichment. Established prognostic tools for kidney allograft failure derive their histological input from a single biopsy at a single time point,^29^ which is precisely the information that a diagnostic history extends.

Several limitations apply. First, treatment was neither randomized nor recorded in detail. We could therefore not establish whether the excess risk of graft failure after a repeated diagnosis arises mainly from suboptimal drug exposure, through inadequate regimens, non-adherence or rapid metabolism, or from the absence of adequate therapeutic options. Either way, current standard practice did not fully eliminate that excess risk. Second, despite adjustment of the graft failure models for pre-transplant HLA-DSA and baseline recipient, donor and transplant characteristics, residual confounding by unmeasured heterogeneity in baseline and time-dependent alloimmune risk between patients may remain. Third, disease may have occurred without being captured in a biopsy, which would bias the estimated effects toward the null. Fourth, the recurrent-event estimates for diagnoses are unadjusted and purely descriptive: associations between different diagnoses partly reflect shared risk and biopsy intensity. Follow-up was also limited to the five-year biopsy period. Fifth, missing values were not imputed, but the small number of transplants excluded for missing data or insufficient follow-up (N=73) had baseline characteristics closely matching the analysis cohort (**Table 1**). Our center also contributed to a recent multicenter study of early TCMR,^7^ so the two cohorts partly overlap for transplants performed between 2013 and 2019. Finally, this is a single-center study, and the protocol biopsy density that makes the central analysis possible is also the principal constraint on its generalizability: the diagnostic trajectories reconstructed here are more complete than those available in most transplant programs, and findings should be extrapolated to centers without systematic protocol biopsies with corresponding caution.

Transition rates into diagnoses were associated with earlier diagnoses, especially but not exclusively those of the same type. A second diagnosis of rejection-related injury was strongly associated with graft failure, and substantially more so than a first, and both remained associated with graft failure when identified on protocol biopsy in patients without graft dysfunction. Even when treated, an episode leaves the graft at a higher risk than before it.

More broadly, these observations indicate that alloimmune injury often unfolds along trajectories, and that longitudinal context is essential for its interpretation and for risk stratification.

## Data Availability

All data produced in the present study are available upon reasonable request to the authors

## Data sharing statement

The data needed to reproduce the findings of this study are not openly available due to reasons of privacy sensitivity and are available from the corresponding author, MN, on reasonable request. Data are located in controlled access data storage at KU Leuven. Analysis code and simulated data will be made available upon publication of the manuscript for the purpose of reproducibility.

## Author contributions

AV, MC and MN designed the study. TD, PK, AP, ThoV, KW, ThiV were involved in data collection and curation. AV performed all analyses with input from MC and MN. AV and MN wrote the manuscript. All co-authors revised the manuscript.

## Supplementary Materials

### Supplementary methods

**Supplementary Table S1.** STROBE checklist

**Supplementary Table S2.** Characteristics of analysis cohort time intervals.

**Supplementary Table S3.** Sequences of diagnoses (N=1562 transplants).

**Supplementary Table S4.** Results of Andersen-Gill models for associations of prior diagnoses with transition rates into later diagnoses (N=1818 transplants).

**Supplementary Table S5.** Sensitivity analysis: results of Andersen-Gill models for associations of prior steroid-treated diagnoses with transition rates into later diagnoses (N=1818 transplants).

**Supplementary Table S6**. Associations of rejection-related injury with the cause-specific hazard of graft failure (N=1818 transplants).

**Supplementary Table S7**. Associations of steroid-treated rejection-related injury with the cause-specific hazard of graft failure (sensitivity analysis, N=1746 transplants).

**Supplementary Table S8.** Associations of specific diagnoses with the cause-specific hazard of graft failure (N=1818 transplants).

**Supplementary Table S9**. Contrast tests to compare second versus first diagnosis of rejection-related injury in terms of their association with the hazard of graft failure.

**Supplementary Figure S1.** Rate ratios of transitions into later diagnoses after steroid-treated earlier diagnoses (sensitivity analysis, N=1818 transplants).

**Supplementary Figure S2.** Associations of sequences of diagnoses with the cause-specific hazard of graft failure (N=1818 transplants).

Supplementary Figure S3. Time-dependent cause-specific hazard ratios for graft failure after a first and a second rejection-related injury (N=1818 transplants).

