## Supplementary Material for "Repeated histological diagnoses and kidney graft failure: an observational cohort study"

**Supplementary methods**

**Supplementary Table S1.** STROBE checklist

**Supplementary Table S2.** Characteristics of analysis cohort time intervals.

**Supplementary Table S3.** Sequences of diagnoses (N=1562 transplants).

**Supplementary Table S4.** Results of Andersen-Gill models for associations of prior diagnoses with transition rates into later diagnoses (N=1818 transplants).

**Supplementary Table S5.** Sensitivity analysis: results of Andersen-Gill models for associations of prior steroid-treated diagnoses with transition rates into later diagnoses (N=1818 transplants).

**Supplementary Table S6.** Associations of rejection-related injury with the cause-specific hazard of graft failure (N=1818 transplants).

**Supplementary Table S7.** Associations of steroid-treated rejection-related injury with the cause-specific hazard of graft failure (sensitivity analysis, N=1746 transplants).

**Supplementary Table S8.** Associations of specific diagnoses with the cause-specific hazard of graft failure (N=1818 transplants).

**Supplementary Table S9.** Contrast tests to compare second versus first diagnosis of rejection-related injury in terms of their association with the hazard of graft failure.

**Supplementary Figure S1.** Rate ratios of transitions into later diagnoses after steroid-treated earlier diagnoses (sensitivity analysis, N=1818 transplants).

**Supplementary Figure S2.** Associations of sequences of diagnoses with the cause-specific hazard of graft failure (N=1818 transplants).

Supplementary Figure S3. Time-dependent cause-specific hazard ratios for graft failure after a first and a second rejection-related injury (N=1818 transplants).

### Supplementary methods

#### Kidney transplant biopsy evaluation

Biopsies of insufficient quality, where the pathologist was not able to assess a single Banff lesion, were not included. Diagnoses were derived from Banff lesions scored by local kidney transplant pathologists and donor-specific anti-human leukocyte antigen antibody (HLA-DSA) status at the time of biopsy. HLA-DSA positive status was defined as a median fluorescence intensity (MFI) above 500, using a Luminex single antigen beads (SAB) assay.

#### Recurrent-event analyses of diagnoses

Transitions between diagnoses were analyzed as recurrent events: each of the six diagnoses can be observed repeatedly and together with other diagnoses. Cumulative incidences of transitions into later diagnoses (Figure 4 of the main text) were estimated from day 90 after transplantation with landmark Aalen-Johansen estimators (mstate R package), separately for the transplants with each diagnosis by day 90, that is, in a biopsy within the first 90 days. The event of interest was the first biopsy after day 90 showing the later diagnosis, graft failure and death with a functioning graft were competing events, and follow-up was censored at five years after transplantation. A transplant with several diagnoses by day 90 contributes to several groups. Curves are shown up to two years after day 90 and stop once fewer than 10 transplants remain at risk.

To summarize the connection between an earlier and a later diagnosis in a single number, transition rates into each diagnosis were modeled with the Andersen-Gill model, an extension of the Cox model for recurrent events. For each pair of an earlier and a later diagnosis, an Andersen-Gill model used non-overlapping start-stop follow-up intervals and an irreversible time-dependent indicator of the earlier diagnosis. The follow-up time does not reset after each event. The Andersen-Gill model uses a jackknife-based robust variance estimator to accommodate dependency between observations. Follow-up was censored at death, at last follow-up, at five years after transplantation, the period in which biopsies were collected, or at the end date of the study (May 2022), whichever came first. Repeated diagnoses in the first 90 days were not counted to minimize biases due to the high incidence of indication biopsies in this timeframe and analyses started at 90 days post-transplantation to avoid immortal time bias. For each follow-up interval, the time-dependent covariate indicates whether the earlier diagnosis had already occurred before or at the start of the time interval. This treats concomitant diagnoses as the simultaneous observation of two separate phenotypes, rather than as a separate phenotype in itself. With six diagnoses, this gave 36 models, each with this single covariate and no adjustment. Like the cumulative incidences, the resulting rate ratios are unadjusted and purely descriptive: they quantify how often one diagnosis follows another, and associations between different diagnoses partly reflect shared risk factors and the intensity of biopsy follow-up. The models included the 1818 transplants with complete covariates, the same transplants as the graft failure models. In a sensitivity analysis, the models were repeated for steroid-treated diagnoses, in 25 models for the five diagnoses other than BKPyVAN. Each of these models contained separate indicators for the steroid-treated and the untreated earlier diagnosis, so that the rate ratio compares a steroid-treated earlier diagnosis with no history of that diagnosis.

#### Multistate model of rejection-related injury and graft failure

The composite exposure, rejection-related injury (RRI), comprised TCMR, AMR and MVI_DSA-/C4d-_. The specific choice of diagnoses for the composite reflects clinical coherence rather than the need for a composite as such: TCMR, AMR and MVI_DSA-/C4d-_ each reflect active injury of established or suspected alloimmune origin and are each managed by escalating immunosuppression. BKPyVAN was analyzed separately throughout: it is a viral infection rather than an alloimmune process and is managed by reducing rather than escalating immunosuppression, so including it in the composite would have produced an exposure with no single coherent treatment implication. bTCMR and pAMR, the borderline and probable variants of TCMR and AMR, were likewise excluded, reflecting their lower diagnostic certainty relative to the definite diagnoses. Their individual associations with graft failure are reported separately in the exploratory analysis.

The multistate model of Figure 1 of the main text has three transient states (no, a first, and a second or later rejection-related injury) and two absorbing states (graft failure and death with a functioning graft). Cause-specific Cox models for the transitions into graft failure used start-stop data, with rejection history updated at each biopsy. The primary exposure had three levels: no rejection-related injury, one rejection-related injury and at least two episodes of rejection-related injury. In order to model long-term associations, these covariates were considered irreversible, meaning that they are carried forward regardless of subsequent No Rejection/No BKPyVAN biopsies. The outcome of the models was the right-censored time from day 90 after transplantation to kidney graft failure, defined as a return to dialysis or retransplantation, or time to end of follow-up, whichever was shorter. Follow-up was censored at death, at last follow-up, at five years after transplantation, or at the end date of the study (May 2022), whichever came first. Repeated diagnoses in the first 90 days were not counted and the analyses started at 90 days post-transplantation, analogously to the Andersen-Gill models. Models were adjusted for pre-transplant HLA-DSA, repeat transplantation, donor type, recipient and donor sex and age, recipient ethnicity and body mass index, cold ischemia time and transplantation year. Wald contrasts compared the coefficients for the second and first rejection-related injury states. Cumulative incidences of graft failure (Figure 6A of the main text) were estimated with the same landmark Aalen-Johansen estimator as Figure 4, from one year after transplantation, separately for the transplants in each rejection-related injury state at one year. Death with a functioning graft was a competing event, and transitions from a first to a second rejection-related injury after one year were allowed. Unlike the classical Aalen-Johansen estimator for multistate models, the landmark estimator does not assume that the process is Markov. A secondary model considered protocol biopsies only, to assess the effects of diagnoses when there is no graft dysfunction. A sensitivity analysis estimated the effects of steroid-treated diagnoses specifically (in the absence of diagnoses without steroid treatment), to verify that results are not solely driven by diagnoses that were not steroid-treated.

HLA-DSA entered all graft failure models as pre-transplant status rather than as a time-dependent covariate. De novo HLA-DSA can arise as a consequence of rejection and in turn drives later injury and graft failure, so that adjusting for its time-dependent status would remove part of the effect under study. Current or historic HLA-DSA also enters the definitions of AMR, pAMR and MVI_DSA-/C4d-_, which require its presence or its absence. Pre-transplant HLA-DSA precedes all post-transplant diagnoses and was therefore treated as a confounder.

Because the proportional hazards test for rejection-related injury in the primary model was borderline non-significant (P = 0.061, Supplementary Table S6), a further model allowed the log hazard ratios of a first and a second rejection-related injury to change linearly with years since transplantation, with the same adjustment set as the primary model. Follow-up intervals were split at each graft failure time, and robust variances clustered by transplant were used. The hazard ratio changed by a factor of 0.68 (95% CI 0.48–0.96) per year for a first and 0.95 (95% CI 0.70–1.29) per year for a second rejection-related injury, so that only the hazard ratio of a first rejection-related injury declined significantly over time. Hazard ratios over time are shown in Supplementary Figure S3.

#### Exploratory analyses and missing data

Separate cause-specific Cox models were fitted for the six individual diagnoses and for each ordered two-diagnosis sequence. Time-dependent covariates for the diagnoses or diagnostic sequences switched permanently from 0 to 1 when the relevant diagnosis or sequence was first observed. These analyses were considered exploratory. Analyses were restricted to complete cases, and no missing values were imputed.

### Supplementary Tables

Supplementary Table S1. STROBE checklist

|  | **Item #** | **Recommendation** | **Page(s)** |
| --- | --- | --- | --- |
| **Title and abstract** | 1 | (*a*) Indicate the study’s design with a commonly used term in the title or the abstract | 1, 3 |
|  |  | (*b*) Provide in the abstract an informative and balanced summary of what was done and what was found | 3 |
| **Introduction** | | | |
| Background/rationale | 2 | Explain the scientific background and rationale for the investigation being reported | 5 |
| Objectives | 3 | State specific objectives, including any prespecified hypotheses | 6 |
| **Methods** | | | |
| Study design | 4 | Present key elements of study design early in the paper | 6 |
| Setting | 5 | Describe the setting, locations, and relevant dates, including periods of recruitment, exposure, follow-up, and data collection | 6, 9 |
| Participants | 6 | (*a*) Give the eligibility criteria, and the sources and methods of selection of participants. Describe methods of follow-up | 6, 9 |
| Variables | 7 | Clearly define all outcomes, exposures, predictors, potential confounders, and effect modifiers. Give diagnostic criteria, if applicable | 7-8 |
| Data sources/ measurement | 8 | For each variable of interest, give sources of data and details of methods of assessment (measurement). | 6-7 |
| Bias | 9 | Describe any efforts to address potential sources of bias | 8, 11-12 |
| Study size | 10 | Explain how the study size was arrived at (if applicable) | NA |
| Quantitative variables | 11 | Explain how quantitative variables were handled in the analyses. If applicable, describe which groupings were chosen and why | 7-8 |
| Statistical methods | 12 | (*a*) Describe all statistical methods, including those used to control for confounding | 7-8 |
|  |  | (*b*) Describe any methods used to examine subgroups and interactions | 7-8 |
|  |  | (*c*) Explain how missing data were addressed | 13 |
|  |  | (*d*) If applicable, explain how loss to follow-up was addressed | 7-8 |
|  |  | (*e*) Describe any sensitivity analyses | 7-8 |
| **Results** | | | |
| Participants | 13 | (a) Report numbers of individuals at each stage of study—eg numbers potentially eligible, examined for eligibility, confirmed eligible, included in the study, completing follow-up, and analyzed | 9, 20 |
|  |  | (b) Give reasons for non-participation at each stage | 9, 20 |
|  |  | (c) Use of a flow diagram | 20 |
| Descriptive data | 14 | (a) Give characteristics of study participants (eg demographic, clinical, social) and information on exposures and potential confounders | 9, 18 |
|  |  | (b) Indicate number of participants with missing data for each variable of interest | 9, 13 |
|  |  | (c) Summarise follow-up time (eg, average and total amount) | 9, 18 |
| Outcome data | 15 | Report numbers of outcome events or summary measures over time | 9, 18 |
| Main results | 16 | (*a*) Give unadjusted estimates and, if applicable, confounder-adjusted estimates and their precision (eg, 95% confidence interval). Make clear which confounders were adjusted for and why they were included | 9-11, 19, 23-24 |
|  |  | (b) Report category boundaries when continuous variables were categorized | NA |
|  |  | (c) If relevant, consider translating estimates of relative risk into absolute risk for a meaningful time period | 9-10, 22, 24 |
| Other analyses | 17 | Report other analyses done—eg analyses of subgroups and interactions, and sensitivity analyses | 10-11 |
| **Discussion** | | | |
| Key results | 18 | Summarise key results with reference to study objectives | 11 |
| Limitations | 19 | Discuss limitations of the study, taking into account sources of potential bias or imprecision. Discuss both direction and magnitude of any potential bias | 13-14 |
| Interpretation | 20 | Give a cautious overall interpretation of results considering objectives, limitations, multiplicity of analyses, results from similar studies, and other relevant evidence | 11-14 |
| Generalizability | 21 | Discuss the generalizability (external validity) of the study results | 14 |
| Funding | 22 | Give the source of funding and the role of the funders for the present study and, if applicable, for the original study on which the present article is based | 14 |

**Supplementary Table S2. Characteristics of analysis cohort time intervals.**

| **Time interval characteristics** | | **Analysis cohort (N=5900)** |
| --- | --- | --- |
| Timing: | |  |
|  | Start time in 1^st^ year since transplantation, no. (%) | 4306 (73.0) |
|  | Start time in 2^nd^ year since transplantation, no. (%) | 1117 (18.9) |
|  | Start time in 3^rd^ year since transplantation, no. (%) | 311 (5.3) |
|  | Start time in 4^th^ year since transplantation, no. (%) | 75 (1.3) |
|  | Start time in 5^th^ year since transplantation, no. (%) | 91 (1.5) |
|  | Start time before 1^st^ biopsy, no. (%) | 682 (11.6) |
|  | Start time at protocol biopsy, no. (%) | 4277 (72.5) |
|  | Start time at indication biopsy, no. (%) | 941 (15.9) |
| Current or historic HLA-DSA presence, no. (%) | | 564 (9.6) |
| Prior diagnoses of rejection-related injury: | |  |
|  | No rejection-related injury, no. (%) | 4418 (74.9) |
|  | First rejection-related injury, no. (%) | 1103 (18.7) |
|  | Second rejection-related injury, no. (%) | 379 (6.4) |

Abbreviations: no., number; HLA-DSA, donor-specific anti-human leukocyte antigen antibodies; TCMR, T-cell mediated rejection; AMR, antibody-mediated rejection; MVI_DSA-/C4d-_, DSA-negative C4d-negative microvascular inflammation; BKPyVAN, BK polyomavirus nephropathy.

Supplementary Table S3. Sequences of diagnoses (N=1562 transplants).

| **Earlier diagnosis** | **Later diagnosis** | **Count** |
| --- | --- | --- |
| (b)TCMR | (b)TCMR | 265 |
| (b)TCMR | (p)AMR | 128 |
| (b)TCMR | BKPyVAN | 34 |
| (b)TCMR | MVI_DSA-/C4d-_ | 65 |
| (b)TCMR | No Rejection/No BKPyVAN | 812 |
| (p)AMR | (b)TCMR | 102 |
| (p)AMR | (p)AMR | 268 |
| (p)AMR | BKPyVAN | 22 |
| (p)AMR | MVI_DSA-/C4d-_ | 16 |
| (p)AMR | No Rejection/No BKPyVAN | 166 |
| BKPyVAN | (b)TCMR | 39 |
| BKPyVAN | (p)AMR | 12 |
| BKPyVAN | BKPyVAN | 46 |
| BKPyVAN | MVI_DSA-/C4d-_ | 14 |
| BKPyVAN | No Rejection/No BKPyVAN | 93 |
| MVI_DSA-/C4d-_ | (b)TCMR | 78 |
| MVI_DSA-/C4d-_ | (p)AMR | 3 |
| MVI_DSA-/C4d-_ | BKPyVAN | 10 |
| MVI_DSA-/C4d-_ | MVI_DSA-/C4d-_ | 82 |
| MVI_DSA-/C4d-_ | No Rejection/No BKPyVAN | 199 |
| No Rejection/No BKPyVAN | (b)TCMR | 611 |
| No Rejection/No BKPyVAN | (p)AMR | 245 |
| No Rejection/No BKPyVAN | BKPyVAN | 171 |
| No Rejection/No BKPyVAN | MVI_DSA-/C4d-_ | 181 |
| No Rejection/No BKPyVAN | No Rejection/No BKPyVAN | 4270 |

Abbreviations: TCMR, T-cell mediated rejection; (b)TCMR, Borderline changes or TCMR; AMR, antibody-mediated rejection; (p)AMR, Probable AMR or AMR; MVI_DSA-/C4d-_, DSA-negative C4d-negative microvascular inflammation; BKPyVAN, BK polyomavirus nephropathy.

Pairs of biopsies within the first five years after transplantation of the 1818 analyzed transplants, the later biopsy after day 90. Earlier biopsies include the first 90 days, as does the diagnostic history in all other analyses.

Supplementary Table S4. Results of Andersen-Gill models for associations of prior diagnoses with transition rates into later diagnoses (N=1818 transplants).

|  | **Covariate** | **RR** | **95% CI** | **P value** |
| --- | --- | --- | --- | --- |
| Incidence of TCMR | |  |  |  |
|  | TCMR | 4.16 | (3.14 - 5.50) | <.0001 |
|  | bTCMR | 1.33 | (0.84 - 2.10) | 0.2294 |
|  | AMR | 2.37 | (1.53 - 3.67) | 0.0001 |
|  | pAMR | 1.94 | (0.85 - 4.44) | 0.1179 |
|  | MVI_DSA-/C4d-_ | 2.30 | (1.52 - 3.47) | <.0001 |
|  | BKPyVAN | 2.20 | (1.31 - 3.69) | 0.0030 |
| Incidence of bTCMR | |  |  |  |
|  | TCMR | 1.28 | (0.92 - 1.77) | 0.1407 |
|  | bTCMR | 1.49 | (1.08 - 2.05) | 0.0149 |
|  | AMR | 1.65 | (1.09 - 2.51) | 0.0181 |
|  | pAMR | 0.77 | (0.23 - 2.52) | 0.6639 |
|  | MVI_DSA-/C4d-_ | 1.45 | (0.95 - 2.21) | 0.0858 |
|  | BKPyVAN | 1.05 | (0.58 - 1.90) | 0.8829 |
| Incidence of AMR | |  |  |  |
|  | TCMR | 2.43 | (1.60 - 3.68) | <.0001 |
|  | bTCMR | 1.75 | (1.06 - 2.88) | 0.0292 |
|  | AMR | 23.67 | (17.39 - 32.21) | <.0001 |
|  | pAMR | 6.51 | (3.76 - 11.25) | <.0001 |
|  | MVI_DSA-/C4d-_ | 0.20 | (0.06 - 0.63) | 0.0057 |
|  | BKPyVAN | 0.89 | (0.38 - 2.12) | 0.7955 |
| Incidence of pAMR | |  |  |  |
|  | TCMR | 1.98 | (0.92 - 4.26) | 0.0808 |
|  | bTCMR | 0.46 | (0.17 - 1.29) | 0.1396 |
|  | AMR | 9.81 | (5.37 - 17.93) | <.0001 |
|  | pAMR | 18.53 | (10.82 - 31.73) | <.0001 |
|  | MVI_DSA-/C4d-_ | NA^a^ | NA | NA |
|  | BKPyVAN | 1.04 | (0.33 - 3.32) | 0.9432 |
| Incidence of MVI_DSA-/C4d-_ | |  |  |  |
|  | TCMR | 1.46 | (0.91 - 2.33) | 0.1148 |
|  | bTCMR | 1.51 | (0.92 - 2.48) | 0.1071 |
|  | AMR | 1.06 | (0.55 - 2.03) | 0.8651 |
|  | pAMR | NA^a^ | NA | NA |
|  | MVI_DSA-/C4d-_ | 8.13 | (5.74 - 11.52) | <.0001 |
|  | BKPyVAN | 1.60 | (0.83 - 3.08) | 0.1585 |
| Incidence of BKPyVAN | |  |  |  |
|  | TCMR | 1.15 | (0.66 - 2.00) | 0.6202 |
|  | bTCMR | 0.42 | (0.20 - 0.86) | 0.0183 |
|  | AMR | 1.32 | (0.71 - 2.44) | 0.3759 |
|  | pAMR | 1.07 | (0.25 - 4.60) | 0.9270 |
|  | MVI_DSA-/C4d-_ | 0.89 | (0.42 - 1.87) | 0.7491 |
|  | BKPyVAN | 7.71 | (5.03 - 11.81) | <.0001 |

Abbreviations: RR, rate ratio; CI, confidence interval, TCMR, T-cell mediated rejection; bTCMR, Borderline changes; AMR, antibody-mediated rejection; pAMR, Probable AMR; MVI_DSA-/C4d-_, DSA-negative C4d-negative microvascular inflammation; BKPyVAN, BK polyomavirus nephropathy; NA, non-applicable.
^a^ Insufficient events to be analyzed.
Each rate ratio is obtained from a separate unadjusted model with the earlier diagnosis as its only covariate. Follow-up started at day 90 and was censored at five years after transplantation.

Supplementary Table S5. Sensitivity analysis: results of Andersen-Gill models for associations of prior steroid-treated diagnoses with transition rates into later diagnoses (N=1818 transplants).

|  | **Covariate** | **RR** | **95% CI** | **P value** |
| --- | --- | --- | --- | --- |
| Incidence of TCMR | |  |  |  |
|  | Treated TCMR | 3.84 | (2.90 - 5.10) | <.0001 |
|  | Treated bTCMR | 1.42 | (0.66 - 3.07) | 0.3753 |
|  | Treated AMR | 3.65 | (2.16 - 6.14) | <.0001 |
|  | Treated pAMR | 1.56 | (0.36 - 6.80) | 0.5558 |
|  | Treated MVI_DSA-/C4d-_ | 2.46 | (1.46 - 4.12) | 0.0007 |
| Incidence of bTCMR | |  |  |  |
|  | Treated TCMR | 1.17 | (0.81 - 1.69) | 0.4092 |
|  | Treated bTCMR | 0.83 | (0.41 - 1.68) | 0.5966 |
|  | Treated AMR | 1.56 | (0.94 - 2.60) | 0.0874 |
|  | Treated pAMR | 0.52 | (0.08 - 3.51) | 0.5047 |
|  | Treated MVI_DSA-/C4d-_ | 0.90 | (0.48 - 1.71) | 0.7537 |
| Incidence of AMR | |  |  |  |
|  | Treated TCMR | 2.50 | (1.60 - 3.91) | <.0001 |
|  | Treated bTCMR | 2.36 | (1.05 - 5.28) | 0.0373 |
|  | Treated AMR | 5.42 | (3.34 - 8.77) | <.0001 |
|  | Treated pAMR | 6.02 | (2.22 - 16.36) | 0.0004 |
|  | Treated MVI_DSA-/C4d-_ | 0.49 | (0.16 - 1.51) | 0.2161 |
| Incidence of pAMR | |  |  |  |
|  | Treated TCMR | 2.18 | (1.01 - 4.69) | 0.0462 |
|  | Treated bTCMR | 0.94 | (0.23 - 3.84) | 0.9338 |
|  | Treated AMR | 3.73 | (1.13 - 12.36) | 0.0312 |
|  | Treated pAMR | 6.13 | (2.49 - 15.13) | <.0001 |
|  | Treated MVI_DSA-/C4d-_ | NA | NA | NA |
| Incidence of MVI_DSA-/C4d-_ | |  |  |  |
|  | Treated TCMR | 1.80 | (1.14 - 2.85) | 0.0117 |
|  | Treated bTCMR | 2.18 | (0.94 - 5.04) | 0.0679 |
|  | Treated AMR | 1.93 | (1.01 - 3.71) | 0.0481 |
|  | Treated pAMR | NA | NA | NA |
|  | Treated MVI_DSA-/C4d-_ | 3.80 | (2.52 - 5.73) | <.0001 |

Abbreviations: RR, rate ratio; CI, confidence interval, TCMR, T-cell mediated rejection; bTCMR, Borderline changes; AMR, antibody-mediated rejection; pAMR, Probable AMR; MVI_DSA-/C4d-_, DSA-negative C4d-negative microvascular inflammation; BKPyVAN, BK polyomavirus nephropathy; NA, non-applicable.
^a^ Insufficient events to be analyzed.
Each rate ratio is obtained from a separate unadjusted model containing the steroid-treated and the untreated history of the earlier diagnosis, and compares a steroid-treated earlier diagnosis with no history of that diagnosis. Follow-up started at day 90 and was censored at five years after transplantation.

**Supplementary Table S6. Associations of rejection-related injury with the cause-specific hazard of graft failure (N=1818 transplants).**

|  | **Covariate** | **HR** | **CI** | **P value** | **cox.zph P value** |
| --- | --- | --- | --- | --- | --- |
| All biopsies | |  |  |  |  |
|  | First rejection-related injury | 2.18 | 1.35 - 3.51 | 0.0014 | 0.0613 |
|  | Second rejection-related injury | 9.31 | 5.84 - 14.85 | <.0001 | 0.0613 |
|  | Pre-transplant HLA-DSA | 1.11 | 0.66 - 1.86 | 0.7057 | 0.3724 |
|  | Repeat transplantation | 1.87 | 1.19 - 2.94 | 0.0065 | 0.0449 |
|  | DCD donor | 0.86 | 0.52 - 1.41 | 0.5456 | 0.9503 |
|  | LD donor | 0.74 | 0.27 - 2.03 | 0.5526 | 0.9503 |
|  | Male recipient sex | 0.82 | 0.57 - 1.19 | 0.3021 | 0.9584 |
|  | Male donor sex | 1.25 | 0.86 - 1.81 | 0.2351 | 0.1506 |
|  | Recipient age, per 10 year increase | 1.00 | 0.86 - 1.16 | 0.9975 | 0.0511 |
|  | Donor age, per 10 year increase | 1.17 | 1.01 - 1.35 | 0.0364 | 0.0465 |
|  | Non-European ethnicity | 1.60 | 0.64 - 4.01 | 0.3170 | 0.8640 |
|  | Recipient BMI | 0.98 | 0.95 - 1.02 | 0.4472 | 0.4237 |
|  | CIT (hours) | 1.01 | 0.97 - 1.05 | 0.5851 | 0.9900 |
|  | Calendar year | 1.03 | 0.98 - 1.07 | 0.2478 | 0.3248 |
| Protocol biopsies | |  |  |  |  |
|  | First rejection-related injury | 1.76 | 1.09 - 2.84 | 0.0202 | 0.7704 |
|  | Second rejection-related injury | 2.58 | 1.31 - 5.08 | 0.0060 | 0.7704 |
|  | Pre-transplant HLA-DSA | 1.83 | 1.07 - 3.12 | 0.0264 | 0.2474 |
|  | Repeat transplantation | 1.94 | 1.24 - 3.05 | 0.0037 | 0.0570 |
|  | DCD donor | 0.91 | 0.55 - 1.48 | 0.6948 | 0.9601 |
|  | LD donor | 0.62 | 0.22 - 1.71 | 0.3560 | 0.9601 |
|  | Male recipient sex | 0.82 | 0.57 - 1.19 | 0.3006 | 0.9351 |
|  | Male donor sex | 1.18 | 0.81 - 1.70 | 0.3896 | 0.1423 |
|  | Recipient age, per 10 year increase | 0.96 | 0.83 - 1.13 | 0.6413 | 0.0524 |
|  | Donor age, per 10 year increase | 1.21 | 1.05 - 1.40 | 0.0095 | 0.0531 |
|  | Non-European ethnicity | 1.49 | 0.60 - 3.71 | 0.3956 | 0.7338 |
|  | Recipient BMI | 0.99 | 0.96 - 1.04 | 0.8028 | 0.4444 |
|  | CIT (hours) | 1.00 | 0.97 - 1.05 | 0.8217 | 0.9982 |
|  | Calendar year | 0.98 | 0.94 - 1.03 | 0.4773 | 0.9999 |

Abbreviations: HR, hazard ratio; CI, confidence interval; HLA-DSA, donor-specific anti-human leukocyte antigen antibodies; DCD, deceased donor (cardiac death); LD, living donor; BMI, body mass index; CIT, cold ischemia time; TCMR, T-cell mediated rejection; AMR, antibody-mediated rejection; MVI_DSA-/C4d-_, DSA-negative C4d-negative microvascular inflammation; BKPyVAN, BK polyomavirus nephropathy.
Results are obtained from two separate models, fit on all biopsies and protocol biopsies respectively. First and Second rejection-related injury were time-dependent covariates, while the other covariates, including pre-transplant HLA-DSA, were time-independent. Cox.zph refers to the proportional hazards test. Follow-up was censored at five years after transplantation.

Supplementary Table S7. Associations of steroid-treated rejection-related injury with the cause-specific hazard of graft failure (sensitivity analysis, N=1746 transplants).

|  | **Covariate** | **HR** | **CI** | **P value** |
| --- | --- | --- | --- | --- |
| All biopsies | |  |  |  |
|  | First rejection-related injury | 2.02 | 1.10 - 3.72 | 0.0237 |
|  | Second rejection-related injury | 12.13 | 5.55 - 26.49 | <.0001 |
|  | Pre-transplant HLA-DSA | 1.55 | 0.72 - 3.34 | 0.2602 |
|  | Repeat transplantation | 2.26 | 1.27 - 4.04 | 0.0057 |
|  | DCD donor | 1.09 | 0.59 - 2.01 | 0.7869 |
|  | LD donor | 1.35 | 0.39 - 4.65 | 0.6384 |
|  | Male recipient sex | 0.92 | 0.57 - 1.49 | 0.7374 |
|  | Male donor sex | 1.46 | 0.90 - 2.36 | 0.1281 |
|  | Recipient age, per 10 year increase | 1.15 | 0.94 - 1.41 | 0.1776 |
|  | Donor age, per 10 year increase | 1.25 | 1.03 - 1.51 | 0.0222 |
|  | Non-European ethnicity | 2.35 | 0.83 - 6.61 | 0.1058 |
|  | Recipient BMI | 0.99 | 0.94 - 1.04 | 0.7135 |
|  | CIT (hours) | 1.03 | 0.98 - 1.08 | 0.2582 |
|  | Calendar year | 1.01 | 0.95 - 1.07 | 0.7677 |

Abbreviations: HR, hazard ratio; CI, confidence interval; HLA-DSA, donor-specific anti-human leukocyte antigen antibodies; DCD, deceased donor (cardiac death); LD, living donor; BMI, body mass index; CIT, cold ischemia time; TCMR, T-cell mediated rejection; AMR, antibody-mediated rejection; MVI_DSA-/C4d-_, DSA-negative C4d-negative microvascular inflammation; BKPyVAN, BK polyomavirus nephropathy.
Results are obtained from a single multivariable model. First and Second rejection-related injury were time-dependent covariates, while the other covariates, including pre-transplant HLA-DSA, were time-independent. 695 follow-up intervals after biopsies with diagnoses not followed by steroid treatment were excluded from this analysis. Follow-up was censored at five years after transplantation.

**Supplementary Table S8. Associations of specific diagnoses with the cause-specific hazard of graft failure (N=1818 transplants).**

| **Model** | **HR** | **CI** | **P value** |
| --- | --- | --- | --- |
| TCMR | 3.54 | 2.42 - 5.18 | <.0001 |
| bTCMR | 1.70 | 1.10 - 2.63 | 0.0175 |
| AMR | 4.66 | 2.69 - 8.05 | <.0001 |
| pAMR | 1.32 | 0.60 - 2.93 | 0.4908 |
| MVI_DSA-/C4d-_ | 3.12 | 1.92 - 5.04 | <.0001 |
| BKPyVAN | 3.33 | 2.02 - 5.48 | <.0001 |
| TCMR to TCMR | 8.32 | 5.11 - 13.54 | <.0001 |
| TCMR to bTCMR | 2.59 | 1.04 - 6.46 | 0.0410 |
| TCMR to AMR | 5.52 | 2.70 - 11.28 | <.0001 |
| TCMR to pAMR | 3.20 | 0.93 - 11.04 | 0.0654 |
| TCMR to MVI_DSA-/C4d-_ | 5.08 | 2.03 - 12.70 | 0.0005 |
| TCMR to BKPyVAN | 11.41 | 5.06 - 25.73 | <.0001 |
| bTCMR to TCMR | 4.31 | 1.94 - 9.58 | 0.0003 |
| bTCMR to bTCMR | 4.98 | 2.28 - 10.88 | <.0001 |
| bTCMR to AMR | 4.59 | 1.94 - 10.83 | 0.0005 |
| bTCMR to pAMR | 4.48 | 0.60 - 33.48 | 0.1439 |
| bTCMR to MVI_DSA-/C4d-_ | 6.91 | 2.73 - 17.50 | <.0001 |
| bTCMR to BKPyVAN | 3.89 | 0.92 - 16.39 | 0.0640 |
| AMR to TCMR | 7.00 | 3.36 - 14.60 | <.0001 |
| AMR to bTCMR | 6.45 | 3.00 - 13.85 | <.0001 |
| AMR to AMR | 4.98 | 2.43 - 10.22 | <.0001 |
| AMR to pAMR | 0.79 | 0.18 - 3.38 | 0.7496 |
| AMR to MVI_DSA-/C4d-_ | 7.60 | 2.35 - 24.52 | 0.0007 |
| AMR to BKPyVAN | 3.55 | 1.21 - 10.35 | 0.0207 |
| pAMR to TCMR | 8.30 | 2.78 - 24.78 | 0.0001 |
| pAMR to bTCMR | 4.87 | 0.63 - 37.37 | 0.1280 |
| pAMR to AMR | 2.98 | 1.02 - 8.74 | 0.0466 |
| pAMR to pAMR | 1.67 | 0.38 - 7.23 | 0.4961 |
| pAMR to MVI_DSA-/C4d-_ | NA^a^ | NA | NA |
| pAMR to BKPyVAN | 44.79 | 9.42 - 213.12 | <.0001 |
| MVI_DSA-/C4d-_ to TCMR | 5.51 | 2.22 - 13.68 | 0.0002 |
| MVI_DSA-/C4d-_ to bTCMR | NA^a^ | NA | NA |
| MVI_DSA-/C4d-_ to AMR | NA^a^ | NA | NA |
| MVI_DSA-/C4d-_ to pAMR | NA^a^ | NA | NA |
| MVI_DSA-/C4d-_ to MVI_DSA-/C4d-_ | 4.31 | 1.97 - 9.40 | 0.0002 |
| MVI_DSA-/C4d-_ to BKPyVAN | 5.61 | 1.34 - 23.42 | 0.0180 |
| BKPyVAN to TCMR | 8.94 | 3.56 - 22.44 | <.0001 |
| BKPyVAN to bTCMR | 3.60 | 0.49 - 26.41 | 0.2085 |
| BKPyVAN to AMR | 6.94 | 2.08 - 23.11 | 0.0016 |
| BKPyVAN to pAMR | NA^a^ | NA | NA |
| BKPyVAN to MVI_DSA-/C4d-_ | 8.93 | 2.77 - 28.77 | 0.0002 |
| BKPyVAN to BKPyVAN | 11.84 | 6.03 - 23.25 | <.0001 |

Abbreviations: HR, hazard ratio; CI, confidence interval; TCMR, T-cell mediated rejection; AMR, antibody-mediated rejection; MVI_DSA-/C4d-_, DSA-negative C4d-negative microvascular inflammation; BKPyVAN, BK polyomavirus nephropathy.
^a^ Insufficient events to be analyzed
Each estimate was obtained from a separate model and each model was adjusted for pre-transplant HLA-DSA, repeat transplantation, donor status, male recipient sex, donor sex, recipient age, donor age, recipient ethnicity, recipient BMI, cold ischemia time (CIT) and calendar year of transplantation. Follow-up was censored at five years after transplantation.

Supplementary Table S9. Contrast tests to compare second versus first diagnosis of rejection-related injury in terms of their association with the hazard of graft failure.

| **Contrast** | **HR** | **95% CI** | **P value** |
| --- | --- | --- | --- |
| Second vs First rejection-related injury in all biopsies | 4.27 | 2.58 - 7.09 | <.0001 |
| Second vs First rejection-related injury in protocol biopsy subset | 1.47 | 0.72 - 2.98 | 0.2924 |

Abbreviations: HR, hazard ratio; CI, confidence interval.

**Supplementary Figures**

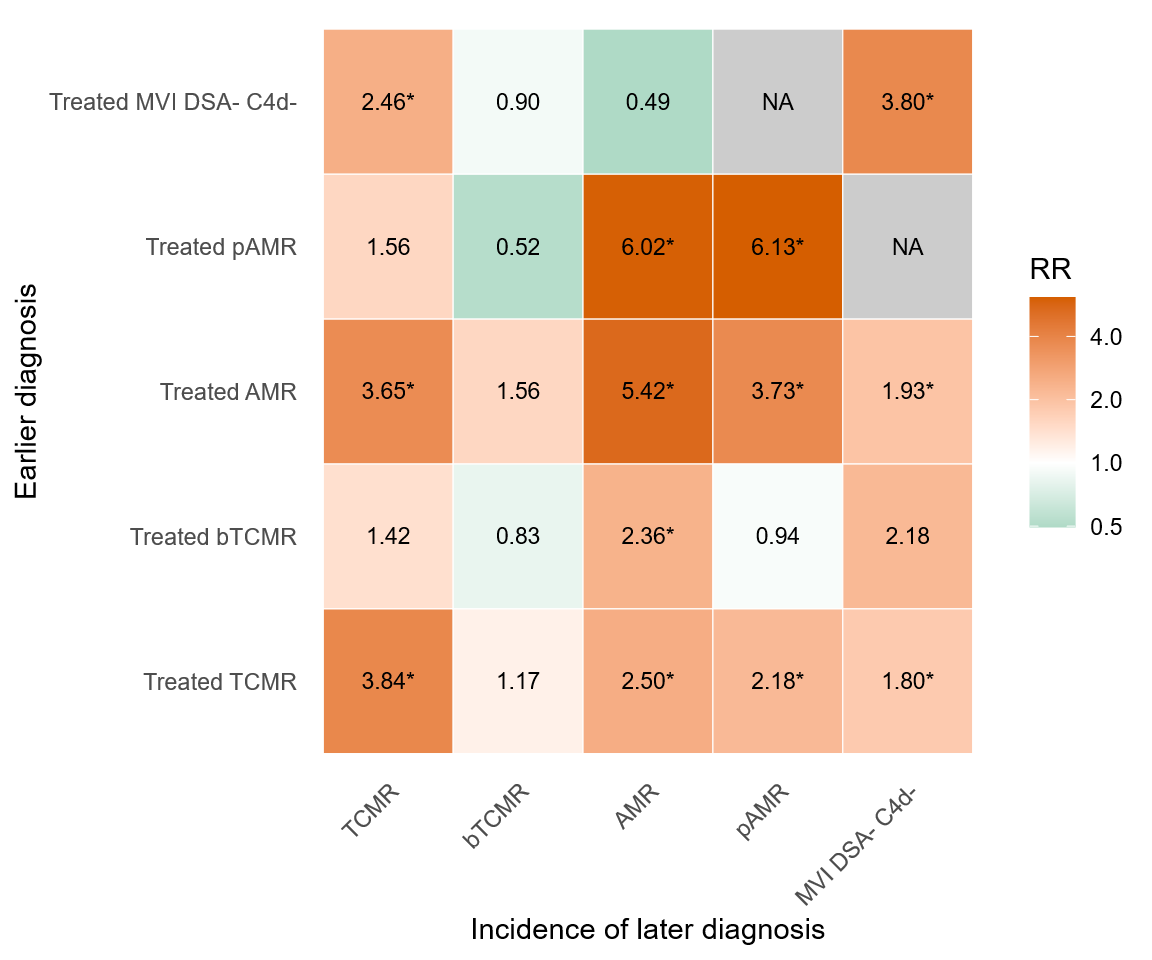

Supplementary **Figure** S1. Rate ratios of transitions into later diagnoses after steroid-treated earlier diagnoses (sensitivity analysis, N=1818 transplants). Each rate ratio is obtained from a separate unadjusted Andersen-Gill model containing the steroid-treated and the untreated history of the earlier diagnosis, and compares a steroid-treated earlier diagnosis with no history of that diagnosis. Follow-up started at day 90 and was censored at five years after transplantation. Abbreviations:RR, rate ratio; NA, non-applicable due to lack of diagnoses; TCMR, T-cell mediated rejection; bTCMR, Borderline changes; AMR, antibody-mediated rejection; pAMR, Probable AMR; MVI_DSA-/C4d-_, DSA-negative C4d-negative microvascular inflammation. *P <.05

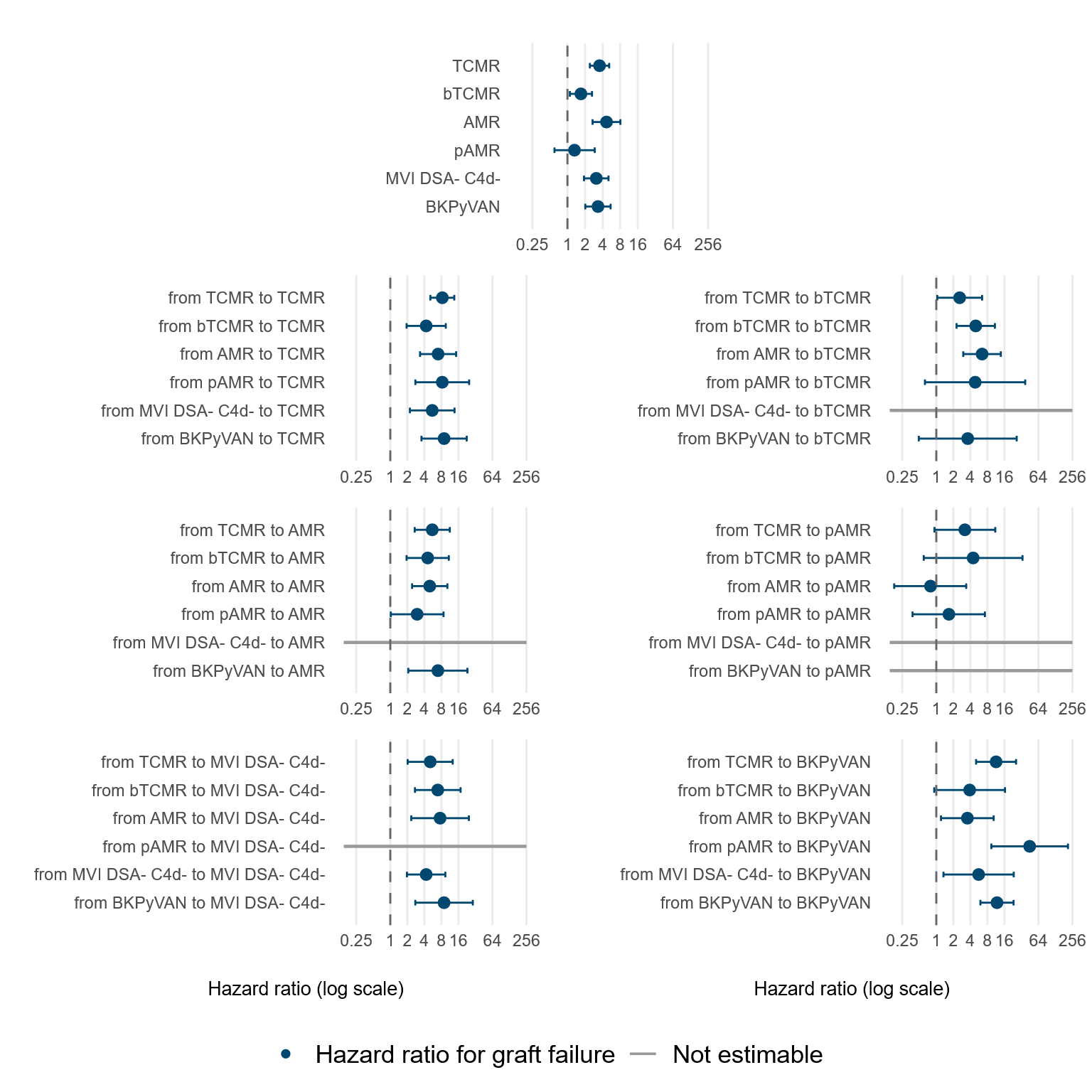

Supplementary Figure S2. Associations of sequences of diagnoses with the cause-specific hazard of graft failure (N=1818 transplants). Every estimate is obtained from a separate model and is adjusted for clinical characteristics at time of transplantation and pre-transplant HLA-DSA status. Follow-up was censored at five years after transplantation. Non-estimable associations were due to insufficient number of graft failure events after some sequences.

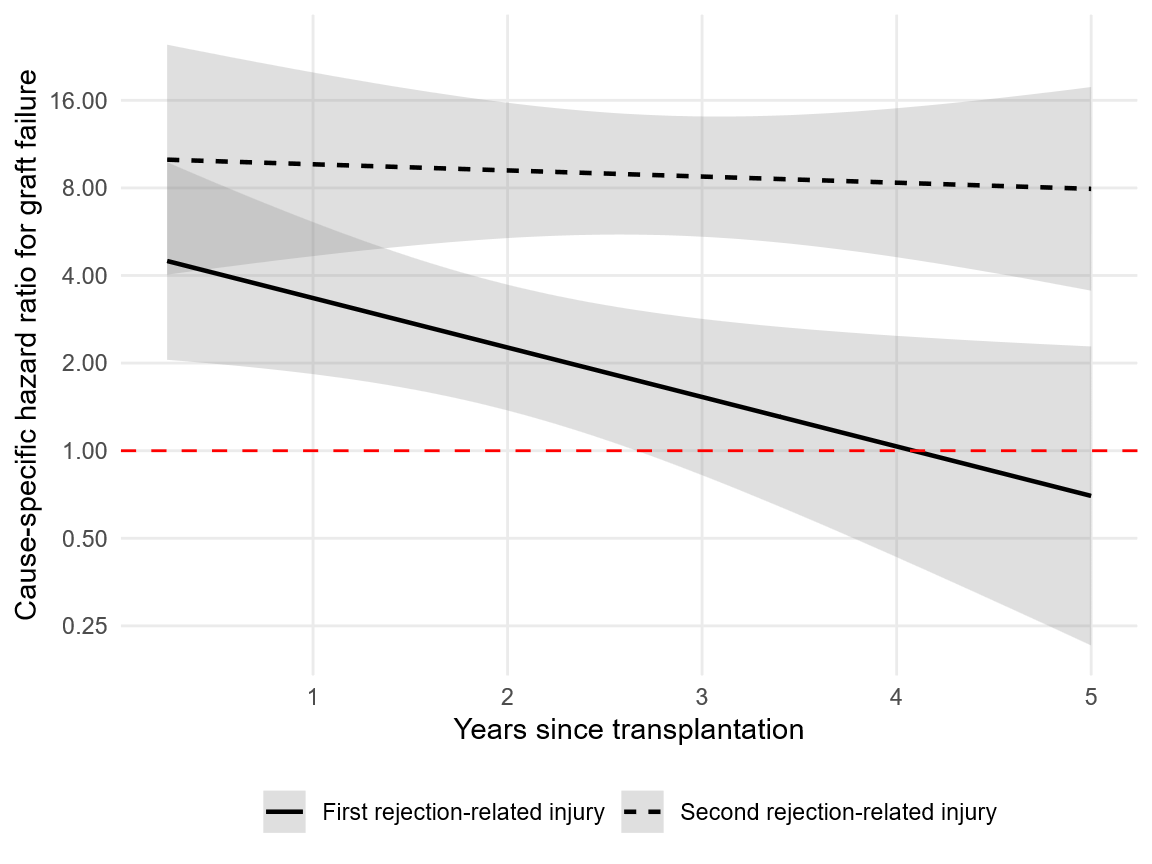

Supplementary Figure S3. Time-dependent cause-specific hazard ratios for graft failure after a first and a second rejection-related injury (N=1818 transplants). The log hazard ratio of each episode was allowed to change linearly with years since transplantation, in a model otherwise identical to the primary model of Supplementary Table S6 and adjusted for clinical characteristics at time of transplantation and pre-transplant HLA-DSA. Shaded bands are 95% confidence intervals and the red dashed line marks a hazard ratio of 1. Follow-up was censored at five years after transplantation.
